# Mosaic deletions in X-linked dystonia-parkinsonism influence repeat stability and disease onset

**DOI:** 10.1101/2022.01.12.22269181

**Authors:** Joanne Trinh, Theresa Lüth, Susen Schaake, Joshua Laβ, Björn-Hergen Laabs, Kathleen Schlüter, Jelena Pozojevic, Ronnie Tse, Inke Köenig, Roland Dominic Jamora, Raymond L. Rosales, Norbert Brüggemann, Gerard Saranza, Cid Czarina E. Diesta, Frank J. Kaiser, Christel Depienne, Ana Westenberger, Christine Klein

## Abstract

**Background:** While multiple genetic causes of movement disorders have been identified in the past decade, modifying factors of disease expression are still largely unknown for most conditions. X-linked dystonia-parkinsonism (XDP) is an inherited neurodegenerative disease caused by a SINE-VNTR-Alu (SVA)-type retrotransposon insertion that contains a hexanucleotide repeat within an intron of the *TAF1* gene. To date, four putative genetic modifiers explain about 65% of variance in age at onset in XDP. However, additional genetic modifiers are conceivably at play in XDP and may include mismatches of the SVA hexanucleotide repeat motif. We aim to identify additional genetic modifiers of XDP expressivity and age at onset (AAO).

**Methods:** Third-generation sequencing of PCR amplicons from XDP patients (n=202) was performed to assess potential repeat interruption and instability. Repeat-primed PCR and Cas9-mediated targeted enrichment were used to confirm the presence of identified repeat mismatches.

**Results:** An increased frequency of deletions at the beginning of the hexanucleotide repeat (*CCCTCT*)_n_ domain was found. Specifically, three deletions at positions 11, 14, and 17 of the *TAF1* SVA repeat motif of somatic mosaic origins were detected in different combinations. The most common one was three deletions (1-2-3) at a median frequency 0.425 (IQR:0.42-0.43) and deletions within positions 11 and 14 (1-2-wt) at a median frequency 0.128 (IQR:0.12-0.13). The frequency of deletions at positions 11 and 14 correlated with repeat number (r=-0.48, p=9.5×10^-13^) and AAO (r=0.34, p=9.5×10^-7^). The association with AAO still stands when including other modifier genotypes (*MSH3* and *PMS2*) in a regression model. However, the association dissipates when including repeat numbers.

**Conclusion:** We present a novel mosaic repeat motif deletion within the hexanucleotide repeat (*CCCTCT*)_n_ domain of *TAF1* SVA. Our study illustrates: 1) the importance of somatic mosaic genotypes; 2) the biological plausibility of multiple modifiers (both germline and somatic) that can have additive effects on repeat instability; 3) that these variations may remain undetected without assessment of single molecules.

## Introduction

While multiple genetic causes of movement disorders have been identified in the past decade, disease modifiers are still largely unknown for most conditions^1^. Individual patients carrying the same pathogenic variant may have variable expressivity of the disease, including variable age at onset (AAO), severity, and clinical manifestations. Thus, in addition to the pathogenic variant, there are genetic modifiers influencing expressivity and onset. One emerging example of this broader concept is X-linked dystonia-parkinsonism (XDP), a neurodegenerative movement disorder endemic to the Philippines and first described in 1976^2,3^.

XDP is characterized by adulthood-onset dystonic movements and parkinsonism due to striatal volume loss as a result of an insertion of the retrotransposon SINE-VNTR-Alu (SVA) in intron 32 of the *TAF1* (TATA-binding protein-associated factor 1) gene^3,4^. There is a hexanucleotide repeat domain within the SVA, which consists of the repeat sequence *(CCCTCT)*_*n*_ ^*5*^. This hexanucleotide repeat domain varies in numbers ranging from 30 to 55 and is a strong genetic modifier of AAO.

To date, there are four putative modifiers of XDP expressivity associated with AAO and/or disease severity^5-7^. These modifiers are the length of the hexanucleotide repeat polymorphism and modifiers of AAO related to DNA mismatch-repair proteins. Both types of modifiers are characteristic for repeat expansion disorders in which expanded repeats of various lengths may be transcribed.

The four putative genetic modifiers originate from germline mutations and, thus, are inherited genetic modifiers. Mosaic modifiers, which exist in every patient at variable frequencies, have not been studied extensively. We previously described the presence of somatic mosaicism in XDP with a higher number of repeats detected in the cerebellum and basal ganglia compared to blood^8^. In this present study, we identified novel mosaic repeat motif patterns that deviate from the known hexanucleotide repeat motif and investigate whether they act as new genetic modifiers of repeat instability and AAO of this neurodegenerative disorder.

## Materials and Methods

### Patient demographics

The study was approved by the Ethics Committees of the University of Lübeck, Germany and at the Metropolitan Medical Center, Manila, Philippines. For the analysis of genomic variants within the SVA and the detection of variations of the hexanucleotide repeat domain, n=202 patients with XDP were investigated and included different brain regions (basal ganglia, cerebellum) and blood-derived DNA from one patient with XDP. As XDP follows an X-linked recessive inheritance pattern, all patients were male. The mean age at onset (AAO) was 41.93 (SD=±8.56) years, and the mean age at examination (AAE) was 47.5 (SD=±9.84) years.

### Nanopore sequencing of PCR amplicon or Cas9 targeted enrichment

All DNA was extracted from the Blood and Cell Culture DNA Midi kit (Qiagen). Sequencing was performed for the SVA amplicon (3.2kb) and Cas9 enrichment as previously described^8^, with super accuracy model for base calling and mapped to reference (detailed in **Supplementary figure 1**). Long-range PCR was performed for the SVA (3.2kb) as previously described1. Subsequently, each patient-derived PCR product was barcoded with the Native 96 Barcoding Kit (EXP-NBD196) and multiplexed. For Cas9 enrichment, crRNAs were designed with ChopChop (https://chopchop.cbu.uib.no). Two crRNAs were used upstream of the TAF1 SVA insertion and two crRNAs were used downstream for a 5.5kb product specifically around the SVA (available upon request). All Libraries were prepared with ligation Sequencing Kit (SQK-LSK109), loaded on a R9.4.1 flow cell and sequenced on MinION/GridION.

### Detection of repeats

Base-calling was performed with the most updated Guppy version 5.0.11. For the detection of the repeat length, the super accuracy model (dna_r9.4.1_450bps_sup.cfg) was used. All reads were mapped to the reference sequence with the software Minimap2 (v2.17) (Supplementary figure 1). Samtools (v1.9) was used for coverage determination and filtering (>1500X). We filtered for Phred score Q>12. Motif mismatch detection was achieved with “Noise-canceling repeat finder” (NCRF) (v1.01.02)^9^. Repeat-primed PCR with a FAM-tagged primer was performed for validation. Primers are available upon request.

### Statistical analyses

Frequencies of deletions were estimated using NCRF and Spearman’s correlation coefficient was employed to estimate the correlation and the corresponding p-values reported. Boxplots were used to show the distribution. Regression models were used to assess the correlation between AAO and the frequency of deletions at position 1 and 2 (1-2-wt), adjusted for three SNPs in MSH3 and PMS2, and adjusted for three SNPs in MSH3 and PMS2 and the SVA repeat number.

### Data availability

The data presented in this study are available on SRA (SAMN24775867-SAMN24775962, SAMN24115523-SAMN24115530). The bioinformatic commands to quantify the TAF1 SVA (*CCCTCT*)n repeat length are described here: https://github.com/nanopol/xdp_sva/.

## Results

Deep sequencing of the PCR amplicon of the 3.2kb *TAF1* SVA region with Nanopore yielded a mean coverage of 10,915X per sample (SD=±8,207) **(Supplementary figure 1)**. We detected a prominent occurrence in every patient (n=202) of deletions with a mean frequency of 0.97 (SD=±0.113) at the beginning of the repeat motif, consistently present on the plus- and minus-strands in all patients **(Figure 1A and 1B)**. At the single nucleotide resolution, three deletions (deletions 1, 2, 3) were found at the 5’ end at positions 11, 14, and 17 of the repeat motif **(Figure 1C)**.

**Figure 1.**
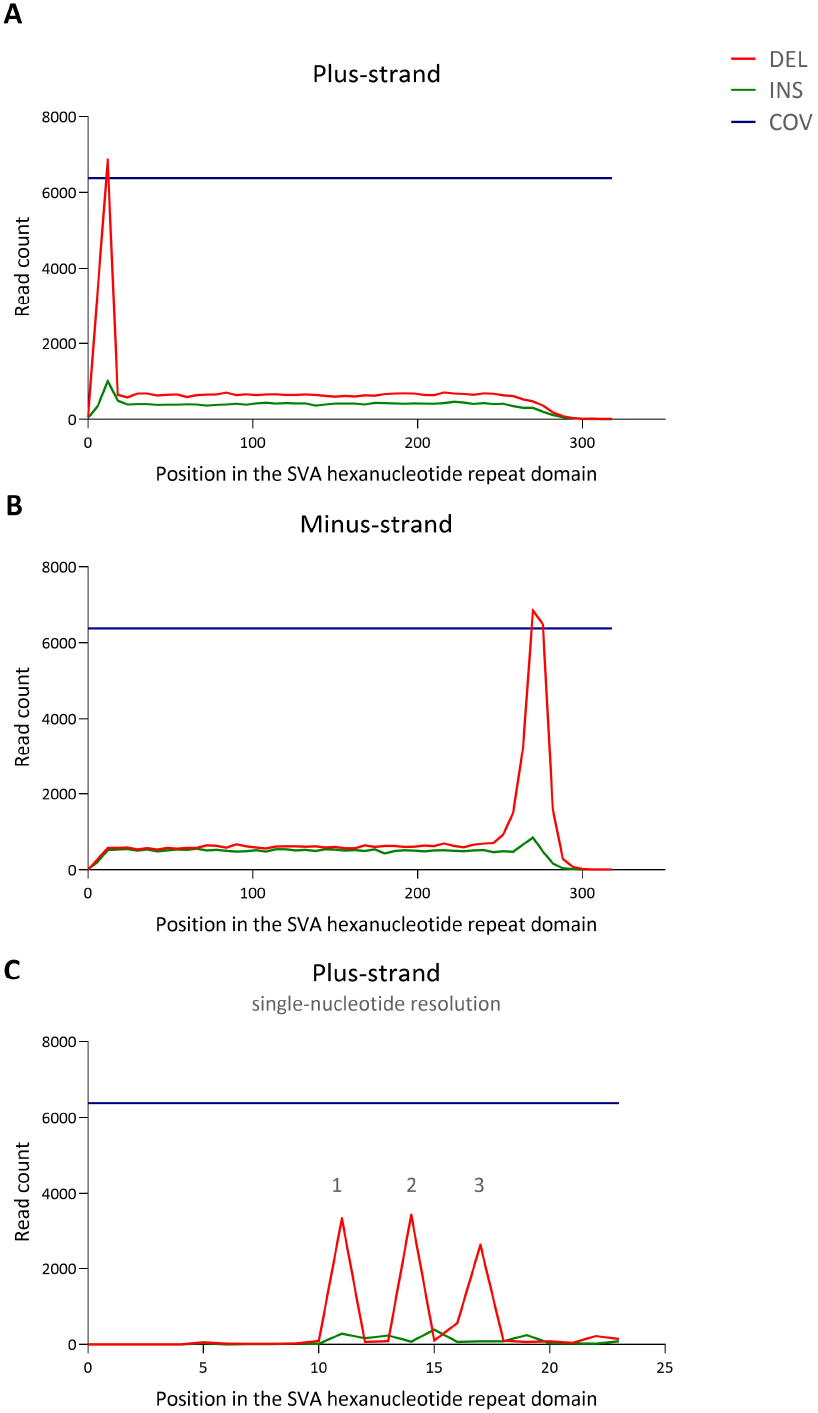
Position of the repeat motif deletion within the *TAF1* SVA hexanucleotide repeat domain. **(A)** Plus-strand reads that have deletions or insertions within the SVA hexanucleotide repeat domain. **(B)** Minus- strand reads that have deletions or insertion within the SVA hexanucleotide repeat domain. **(C)** Single-nucleotide resolution of the deletions detected at the 5’ end of the SVA hexanucleotide repeat domain. Legend: DEL=Deletion, INS=Insertion, COV=Coverage (number of reads covering the *TAF1* SVA hexanucleotide repeat domain)

The deletions were detected at multiple combinations and various frequencies in every patient, indicating somatic mosaicism **(Figure 2A)**. The most frequently detected deletion in all of the analyzed patients in this study was the triple deletion (1-2-3) with the repeat motif pattern (AGAGGG)_2_ AGG (AGAGGG)_n_ and a median frequency 0.425 (IQR:0.42-0.43). The second most detected deletion combination was within position 11 and 14 (1-2-wt), where the resulting repeat motif pattern was (AGAGGG)_2_AGGG(AGAGGG)_n_ and the median frequency 0.128 (IQR:0.12-0.13) **(Figure 2B)**.

**Figure 2.**
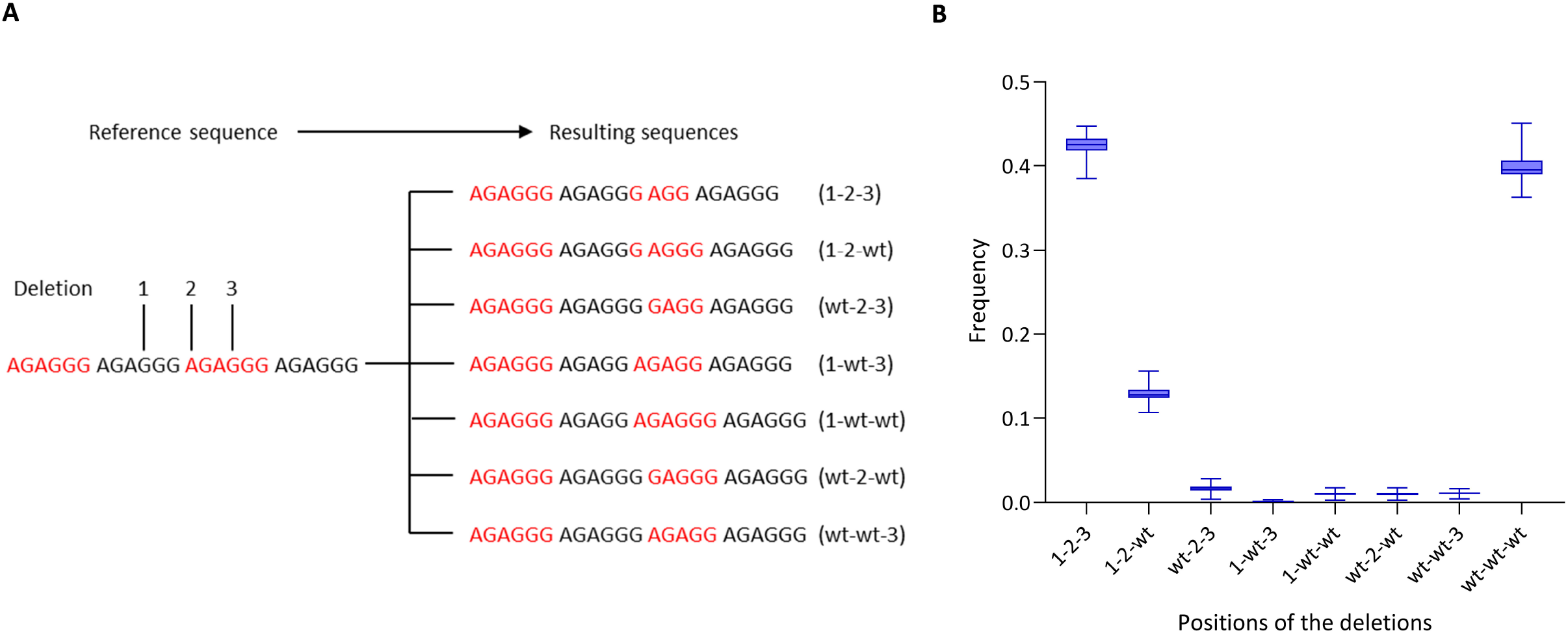
Overview of the deletion combinations within the *TAF1* SVA hexanucleotide repeat motif and the corresponding detected frequencies. **(A)** The positions of detected deletions are displayed alongside the resulting sequences of the interrupted repeat motif. **(B)** The boxplot shows frequencies of the combinations of deletions. The line and box represent median and interquartile range respectively and the whiskers represent the range.

Further genetic validation was performed using repeat-primed PCR and Cas9-targeted enrichment of the deletions. Using repeat-primed PCR targeting the three deletions, we observed a signal that indicated the presence of deletions at the beginning of the repeat motif **(Supplementary figure 2)**. Lastly, Cas9- targeted enrichment was performed to avoid errors from PCR amplification as another validation. We found comparable frequencies of somatic deletions in the blood, basal ganglia, and cerebellum-derived DNA **(Supplementary figure 3)**.

We focused our further analyses on the double-deletion (1-2-wt) and triple-deletion (1-2-3) combinations as they were the most frequently detected. We investigated these deletions and their influence on repeat number. The frequency of the two-deletion haplotype negatively correlated with repeat number (r=-0.48, *p*=9.5×10^-13^): the higher the frequency of 1-2-wt, the lower the number of repeats **(Figure 3A)**. This same effect was not observed for the triple deletions **(Figure 3B)**. The double-deletion haplotype positively correlated with AAO (r=0.34, *p*=9.5×10^-7^), whereas the triple-deletion did not show a correlation with AAO **(Figure 3C and D)**. The association still stands when including other modifier genotypes (tagging SNPs related to *MSH3* and *PMS2*) in a regression model. However, the association dissipates when including repeat numbers **(Supplementary Figure 4A-C)**, which infers that the loss of the deletion indirectly delays the AAO.

**Figure 3.**
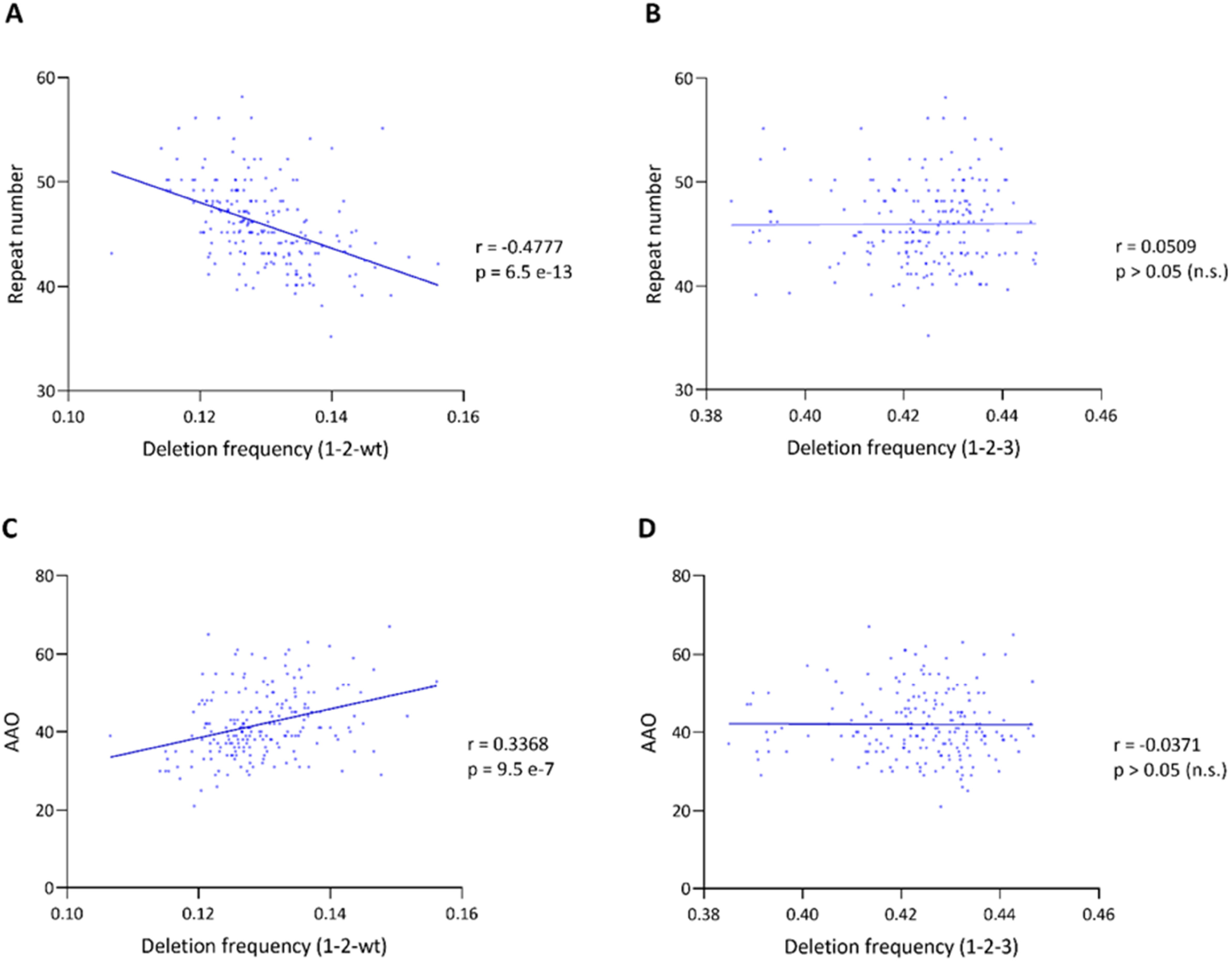
Relationship between repeat motif deletions, age at onset, and repeat number in patients with XDP. **(A)** The correlation between hexanucleotide repeat number and the frequency of long reads with deletions only at positions 1 and 2 (1-2-wt). **(B)** The correlation between hexanucleotide repeat number and deletions at all three positions (1-2-3). **(C)** The correlation between age at onset (AAO) and frequency of deletions only at positions 1 and 2 (1-2-wt). **(D)** The correlation between age at onset (AAO) and frequency of deletions at all three positions (1-2-3). r=Spearman’s rank correlation coefficient, p= Spearman’s exploratory p-value

## Discussion

A thorough analysis of repeat motif mismatches resulted in the detection of deletions in the repeat motif. Repeat mismatches exist in other repeat expansion disorders^10-13^. For example, small interruptions of the repeat motif at the 3’ end of an expansion of the (*GAA*) in Friedreich’s ataxia^14^ delays AAO by nine years. Another example is the loss of interruption at the 3’ end in the *HTT CAG*- repeats, associated with an earlier onset of Huntington’s disease^15^. However, these disruptions in repeats are not reported as somatic and putative mosaic repeat disruptions have not been looked at. XDP deletions were found at the beginning (5’ end) of the *TAF1* SVA repeat motif in a somatic mosaic fashion, indicating a new mechanism. We postulate that these deletions stabilize the repeats, especially as a higher frequency of the 1-2-wt combination is associated with shorter repeats. It can be inferred that the loss of the deletion indirectly delays the AAO.

General limitations of Nanopore sequencing are possible artifacts by amplification during the long-range PCR, sequencing or software^9^. Thus, for further confirmation, we validated the presence of deletions in two ways: RP-PCR and Cas9-targeted enrichment. However, somatic mosaicism is difficult to detect and present only with Cas9 enrichment and PCR amplicon long-read sequencing. Still, replication in other cohorts and using different technologies is warranted.

Our study illustrates: 1) the importance of underexplored somatic mosaic genotypes, present in every individual in this case; 2) the biological plausibility of multiple modifiers (both germline and somatic) that can have effects on repeat instability and expressivity; 3) that these variations may remain undetected with older technologies that do not assess single molecules. Importantly, this study sheds light on another putative modifier of XDP expressivity associated with AAO and potentially disease severity. Mosaic repeat deletions present as a novel disease mechanism that is also clinically relevant for other repeat expansion disorders and future genetic counseling with implications beyond XDP.

## Data Availability

All data produced in the present study are available upon reasonable request to the authors

https://github.com/nanopol/xdp_sva/

## Acknowledgment and Funding

The study was supported by the Deutsche Forschungsgemeinschaft (DFG FOR2488; to JT, IRK, NB, AW, and CK), by funding by the Collaborative Center for X-linked Dystonia-Parkinsonism at Massachusetts General Hospital (to NB and CK), the Else-Kröner Fresenius Foundation (to JT and NB), by intramural funds from the University of Lübeck (to JT and CK), career development award from Peter Engelhorn Foundation (to JT) and by a career development award from the Hermann and Lilly Schilling Foundation (to CK).

The authors declare no conflict of interest. JT and TL had full access to all the data in the study and takes responsibility for the integrity of the data and the accuracy of the data analysis.

## Supplementary material

Supplementary material is available at Brain online

