## Supplementary data for "Mosaic deletions in X-linked dystonia-parkinsonism influence repeat stability and disease onset"

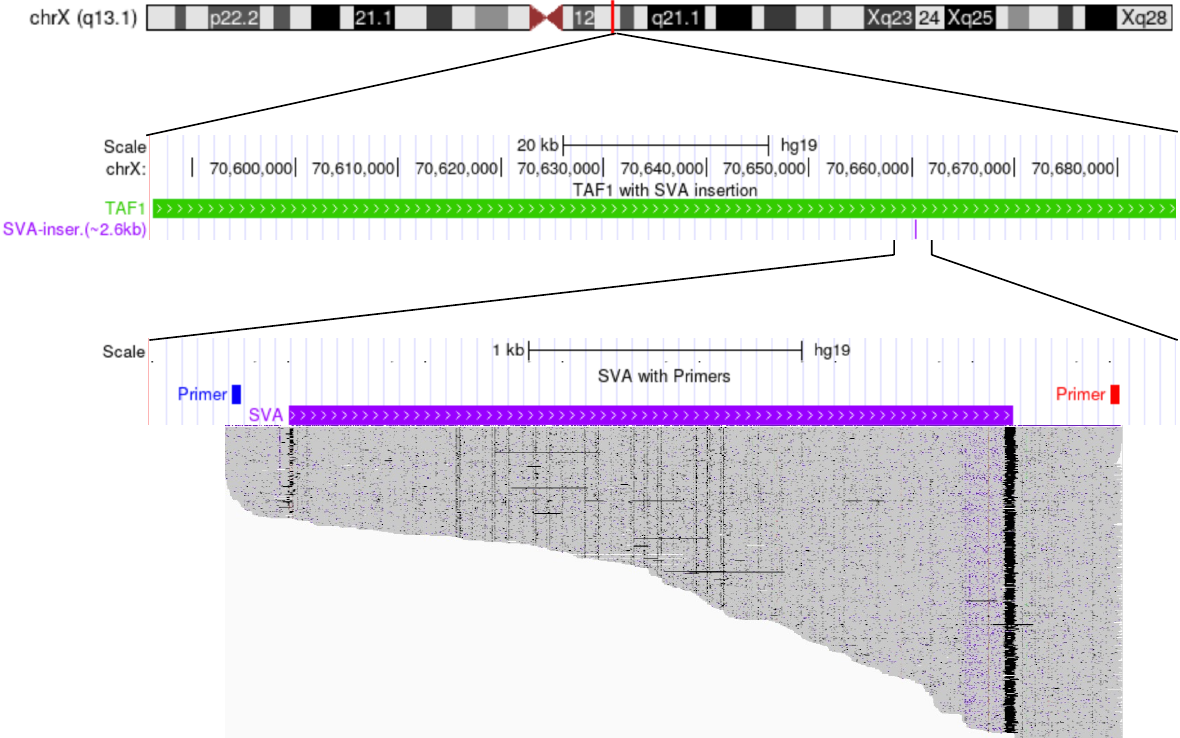

**Supplementary Figure 1. Position of the *TAF1* SVA retrotransposon and aligned long reads.** Genomic location of the *TAF1* gene (green bar) on the X chromosome and the SVA retrotransposon insertion (purple bar) in patients with XDP. The binding site of the primer pair (red and blue bar) is shown, which was used to amplify the *TAF1* SVA. Reads obtained from long-read Nanopore sequencing for one sample are displayed.

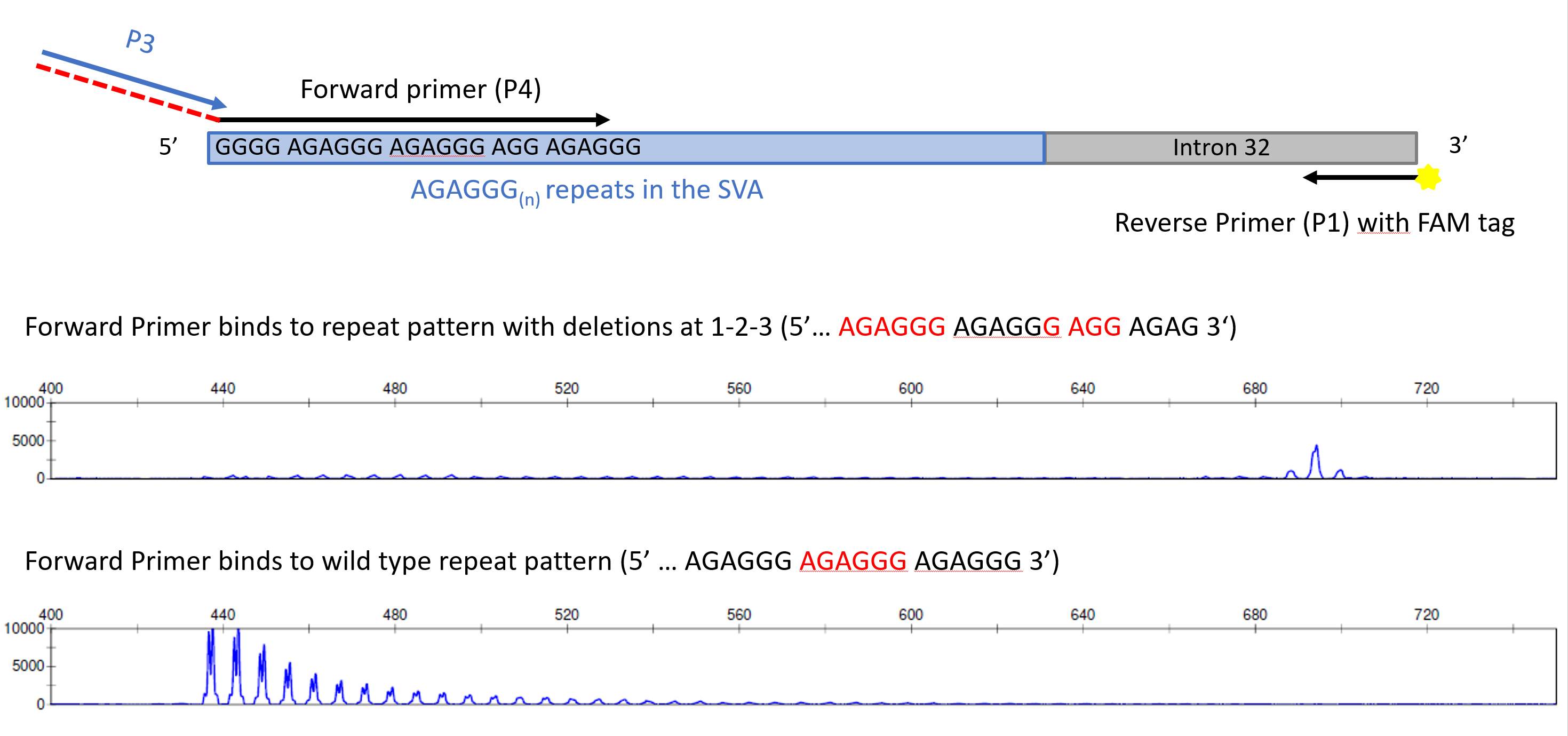

**Supplementary figure 2. Repeat-primed PCR was used to validate the interrupted repeat motif.** Schematic of the primer combinations used for the repeat-primed PCR. The forward primer P4 binds specifically to different interrupted *TAF1* SVA hexanucleotide repeat motifs and has a tail (red dashed line) to which the tail-specific primer P3 binds. The reverse primer P1 is 6-FAM-labelled and the binding site is located within intron 32. Example electropherograms of repeat primed PCR for an XDP patient (L-8311) with a repeat number of n=43. PCR with primer P4 binding to deletion combination 1-2-3, showing a peak around 700 bp; or to the wildtype repeat pattern (wt-wt-wt), showing repeat traces starting at 440 bp.

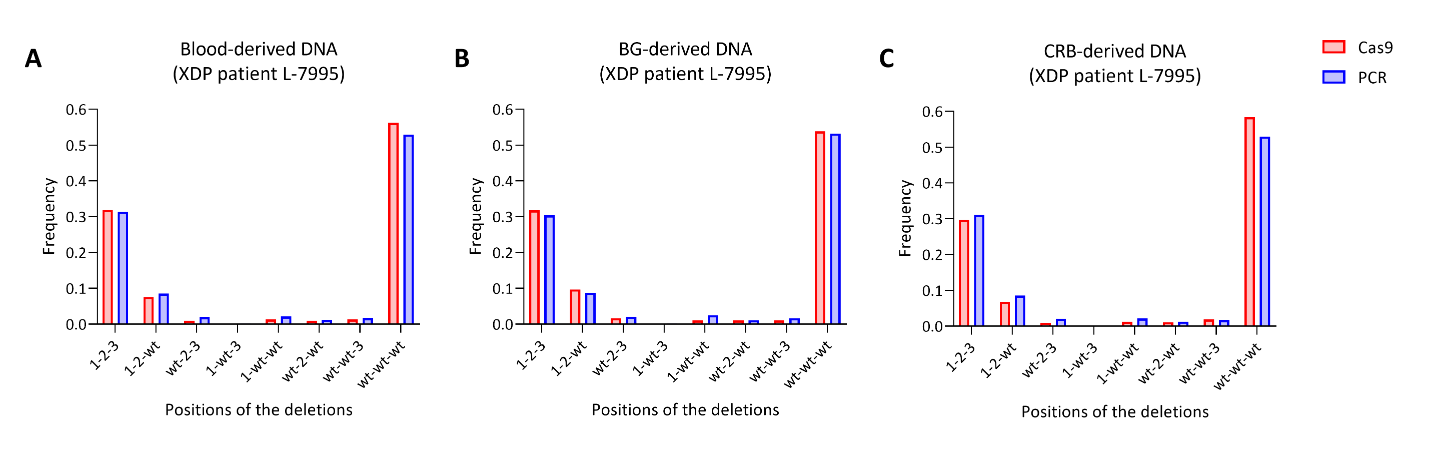

**Supplementary figure 3. Comparison of deletion frequencies detected in blood- and brain-derived DNA, enriched for the SVA insertion by PCR or amplification-free Cas9-enrichment.** The bars represent a single frequency value for each deletion combination detected from **(A)** blood-derived DNA, **(B)** basal ganglia (BG)-derived DNA and **(C)** cerebellum (CRB)-derived DNA. The bar charts show the detected frequencies of the different combinations of deletions.

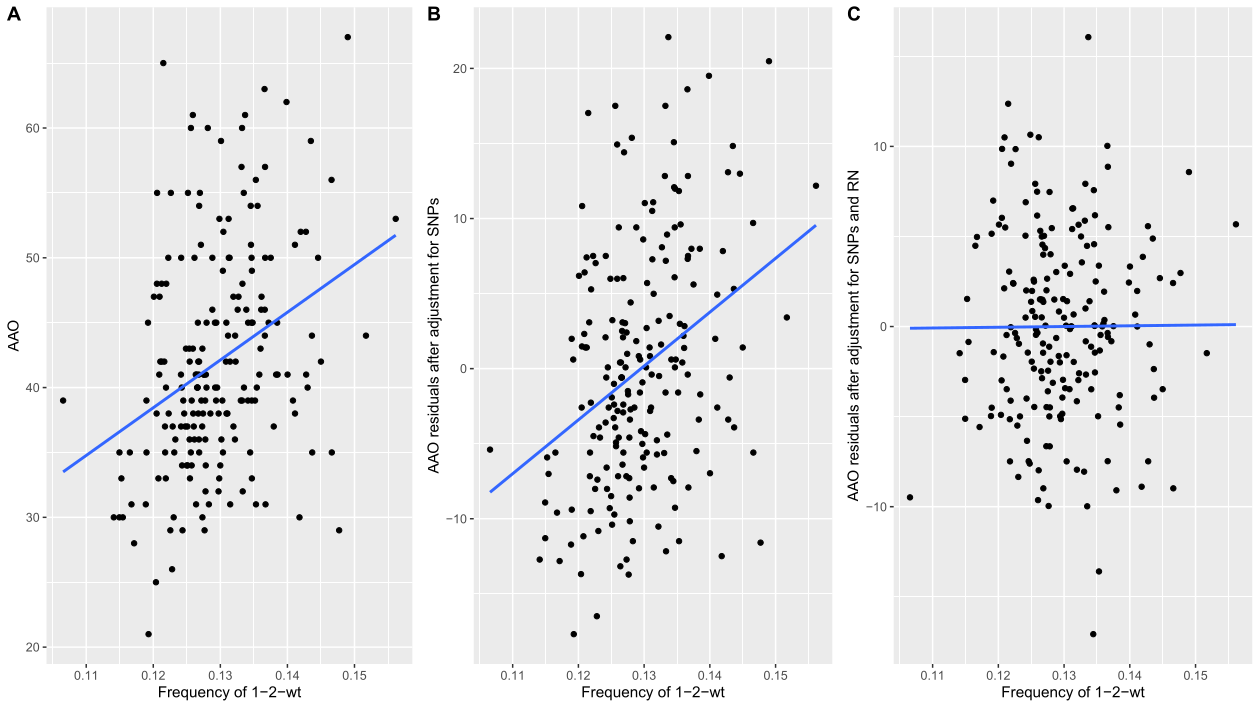

**Supplementary figure 4. Association between age at onset (AAO) and repeat deletions in patients with XDP.** The plots show **(A)** the correlation between AAO and the frequency of deletions at position 1 and 2 (1-2-wt), **(B)** adjusted for three SNPs in *MSH3* and *PMS2*, **(C)** adjusted for three SNPs in *MSH3* and *PMS2* and the SVA repeat number.
